# The Bidirectional Relationship Between Head Injuries and Conduct Problems: Longitudinal Modelling of a Population-Based Birth Cohort Study

**DOI:** 10.1101/2022.11.21.22282449

**Authors:** Hannah R. Carr, James E. Hall, Hedwig Eisenbarth, Valerie C. Brandt

## Abstract

Childhood head injuries and conduct problems increase the risk of aggression and criminality and are well-known correlates. However, the direction and timing of their association and the role of their demographic risk factors remain unclear. This study investigates the bidirectional links between both from 3 to 17 years while revealing common and unique demographic risks.

A total of 7,140 participants (51% female; 83.9% White ethnicity) from the Millennium Cohort Study were analysed at 6 timepoints from age 3 to 17. Conduct problems were parent-reported for ages 3 to 17 using the Strengths and Difficulties Questionnaire (SDQ) and head injuries at ages 3 to 14. A cross-lagged path model estimated the longitudinal bidirectional effects between the two whilst salient demographic risks were modelled cumulatively at three ecological levels (child, mother, and household).

Conduct problems at age 7 promoted head injuries between 7 and 11 (*Z* = .07; *SE* = .03; 95% CI, .01-.12), and head injuries then promoted conduct problems at age 14 (*ß* = .07; *SE* = .03; 95% CI, .01-.12). Head injuries were associated with direct child-level risk until 7 years, whereas conduct problems were associated with direct risks from all ecological levels up until 17 years.

The findings suggest a sensitive period at 7 to 11 years for the bidirectional relationship shared between head injuries and conduct problems. They suggest that demographic risks for increased head injuries play an earlier role than they do for conduct problems. Both findings have implications for intervention timing.

## Introduction

Childhood conduct problems and childhood head injuries are both significant risk factors for lifelong aggression and criminality [1,2] and are known correlates [3]. However, how and when conduct problems and head injuries increase the other during childhood, particularly when controlling for demographic risk factors, remains unknown. This poses a serious problem for professionals in health, social care, and education. Without knowing when and to what extent head injuries pose a risk for conduct problems (and vice versa) it is difficult to design and deploy interventions with the greatest potential for impact.

Conduct problems can be defined as repeated violations to age-appropriate societal norms [4], such as fighting, threatening, and bullying. If these behaviours persist, a conduct disorder diagnosis may be warranted. One of the potential causes of conduct problems is head injury [5]. Head injury is the main cause of death and disability in the UK, with approximately 1.4 million admissions of head injury every year, of which 33% to 55% are children [6].

Clinical studies have shown increased conduct problems following traumatic brain injuries (TBI) [5,7]. Mild head injuries (those that do not disrupt normal brain functioning) are similarly associated with greater levels of conduct problems in adolescence and early adulthood [8], as well as a slower decline in impulsivity and greater levels of reactive aggression by adolescence [2]. Mechanisms explaining how head injuries pose a risk for increased conduct problems include changes to brain areas linked with executive functioning and fear processing,[9] and changes in neural connectivity [10].

There is limited published research investigating whether childhood conduct problems also influence the risk of later head injuries. With studies either investigating this relationship alongside other co-morbid diagnoses such as ADHD [11] or between adolescent conduct problems and adulthood head injuries [12]. However, a recent study suggests that childhood conduct problems can similarly predict an increased risk of sustaining head injuries by age 14 [3].

Although the current literature may suggest a potential bidirectional association between childhood conduct problems and head injury, no published study has explicitly investigated this association, nor identified a sensitive age in which these associations take place. This information is critical to inform effective interventions. Limitations of many previous studies is their focus on TBIs, while 95% of head injuries are mild or never reported [6]. Many also include clinical samples, self-reported head injuries, use long delays in reporting of head injuries, and failure to control for common factors influencing both conduct problems and head injuries. We sought to account for such limitations by investigating whether there is a bidirectional association between head injuries and conduct problems during child development from 3 to 17 years in a large, longitudinal UK cohort. Importantly, the current study controls for salient demographic risk factors concerning the child, their mother, and their household, leading to two research questions:

1. Are there bidirectional associations between head injuries and conduct problems from ages 3 to 17 years?
2. Is combined risk at the child, mother and household levels associated with conduct problems and/or head injuries from ages 3 to 17 years?

## Methods

### Study design and participants

Participants were part of the Millennium Cohort study (MCS), a longitudinal birth cohort study of 18,786 individuals born in the UK between 2000 and 2002 [13]. They were studied at seven time points, at the ages of 9 months (T1), 3 (T2), 5 (T3), 7 (T4), 11 (T5), 14 (T6), and 17 years (T7). Following common practice in longitudinal studies the analyses were limited to those with complete conduct problem data at the last wave (T7) [14]. Further exclusions were made to those with a reported diagnosis of either ADHD, epilepsy, or both as they are associated with an increased risk of head injury [15,16]. Those who were not first-born children were also excluded. There appears to be different levels of aggression related schemas in first, second, and third children [17], as well as higher risks of injuries in first-born children [18], which could influence the levels of conduct problem and head injuries highlighted in this study. Final exclusions were made to those whose main respondent in the study was not their biological mother because risk factors such as mother to child attachment were measured only for the biological mother. If we did not include this exclusion a proportion of the sample would not have available data for all risk factors at the mother-level. This resulted in an analytic sample of 7,140 individuals (3,647 female [51.1%]; 83.9% White ethnicity). See flow chart in Supplementary Figure 1 for breakdown.

All procedures and analyses were approved by the University of Southampton Ethics Committee (ID=62100). Families provided written informed consent to take part and consented for their data to be shared for secondary analysis. Data were downloaded from the UK Data Archive [beta.ukdataservice.ac.uk/datacatalogue/series/series?id=2000031].

## Measures

### Conduct Problems

These were assessed from age 3 (T2) using the five items from the Conduct Problem Subscale of the parent-report version of the Strengths and Difficulties Questionnaire (SDQ) [19]. Items are scored on a 3-point scale (0 - 2) with a higher total score indicating a higher level of conduct problems (possible range: 0 - 10). Cronbach’s alpha values within this study ranged from .52 to .66 across the MCS waves. Previous research has shown the SDQ to have over 75% sensitivity in identifying clinically relevant conduct problems [20] and the parent version specifically has strong validity in identifying conduct disorder [21].

### Head Injuries

Parents were asked if their child had ever had a head injury that resulted in them being taken to the doctor, health centre, or hospital at age 9 months (T1) and at subsequent waves, if their child had sustained any further head injuries since the last wave. Head injuries (coded 1) included responses categorised as a ‘bang on the head’ or ‘loss of consciousness’. The ‘loss of consciousness’ group was extremely small (n ranging from 22 to 85 across waves) meaning that there would not have been the statistical power to warrant analysing the groups separately. The overall ‘head injury’ variables (one per wave) also capture everyday head injuries sustained in the general population as opposed to the moderate-severe head injuries that are often the focus of the literature. Head injury data was only analysed from T2 (3 years) onwards to achieve temporal ordering with the studies risk factors which were measured at T1 and T2.

### Demographic Risks

Demographic risks were divided by ecological level (child, mother, and household) and the combined risk from each level was measured via the construction of a cumulative risk index (CRI). Each CRI consisted of five items which were dichotomised into 0’s (low risk) and 1’s (high risk) based on the literature and then summated. A higher score indicated the presence of more risks in a child’s development (0 - 5 risks present per ecological level). Further details of each CRI can be seen in Supplementary Table 1.

#### Child Level Risk

Child level risk factors were taken from the parent interview at T1 and included male sex [2,22], low birth weight (<2.5 kg) and premature birth (<=252 days gestation) [23,24], and (due to biological effects on the child) whether the child’s biological mother smoked or drank alcohol during pregnancy [25,26].

#### Mother Level Risk

Mother level risk factors were also from the parent interview at T1 and included pregnancy before 18 years [22,27], no high-school qualification [25-28], current unemployment [25,26], low attachment with child (<= 22 on Condon Maternal Attachment Scale) [29,30], and psychological distress (>4 on Rutter Malaise Inventory) [22,31].

Attachment with child was measured using a subset of six items from the Condon Maternal Attachment Questionnaire [29]. The items were scored on a scale from 1 (*almost all the time*) to 5 (*never*; possible range: 0 - 30). A lower score indicates greater difficulties in mother-child attachment. Maternal psychological distress was measured using the MCS’s 9-item composite variable of the Rutter Malaise Inventory’s original 24-item scale [31]. The items included ‘*Are you easily upset or irritated?*’ and were coded as 0 (*no)* and 1 (*yes)* and summed (possible range: 0 – 9) with a higher score indicating higher psychological distress.

#### Household Level Risk

Household level risk factors were taken from the parent interview at T1 and T2. These included single parent household [27,32], low household income (< 60% of median household income) [27,32], household overcrowding (fewer rooms than people excluding bathrooms and hallways) [27,32], low household occupational status (highest occupational status in the household being semi-skilled or lower) [28], and a low-quality home learning environment (bottom quartile of early home learning environment scale) [33]. The home learning environment was measured at T2 using six of the original seven items used in the home learning environment scale available in the MCS dataset (excluding ‘*playing with numbers’*) [33]. These measured the frequency at which the child engaged in learning activities, such as going to the library. These items were scored on a rating-scale from 0 (*not at all*) to 7 (*everyday*) and summed (possible range 0 - 42). A higher score indicates a higher quality home learning environment.

#### Statistical Analysis

Mplus (v7.4) was used to run a cross-lagged path model (see Figure 1) to test the relationships between head injury and conduct problems over time while controlling for salient demographic risks. Sample to UK population generalisation was bolstered through use of the MCS sample weights from T7, which accounted for stratification, attrition, and nonresponse bias. The internal validity of the statistical estimates concerning the binary head injury variables were improved through use of the weighted least square estimation procedure. Missing data were accounted for though use of the Full Information Maximum Likelihood procedure.

**Figure 1.**
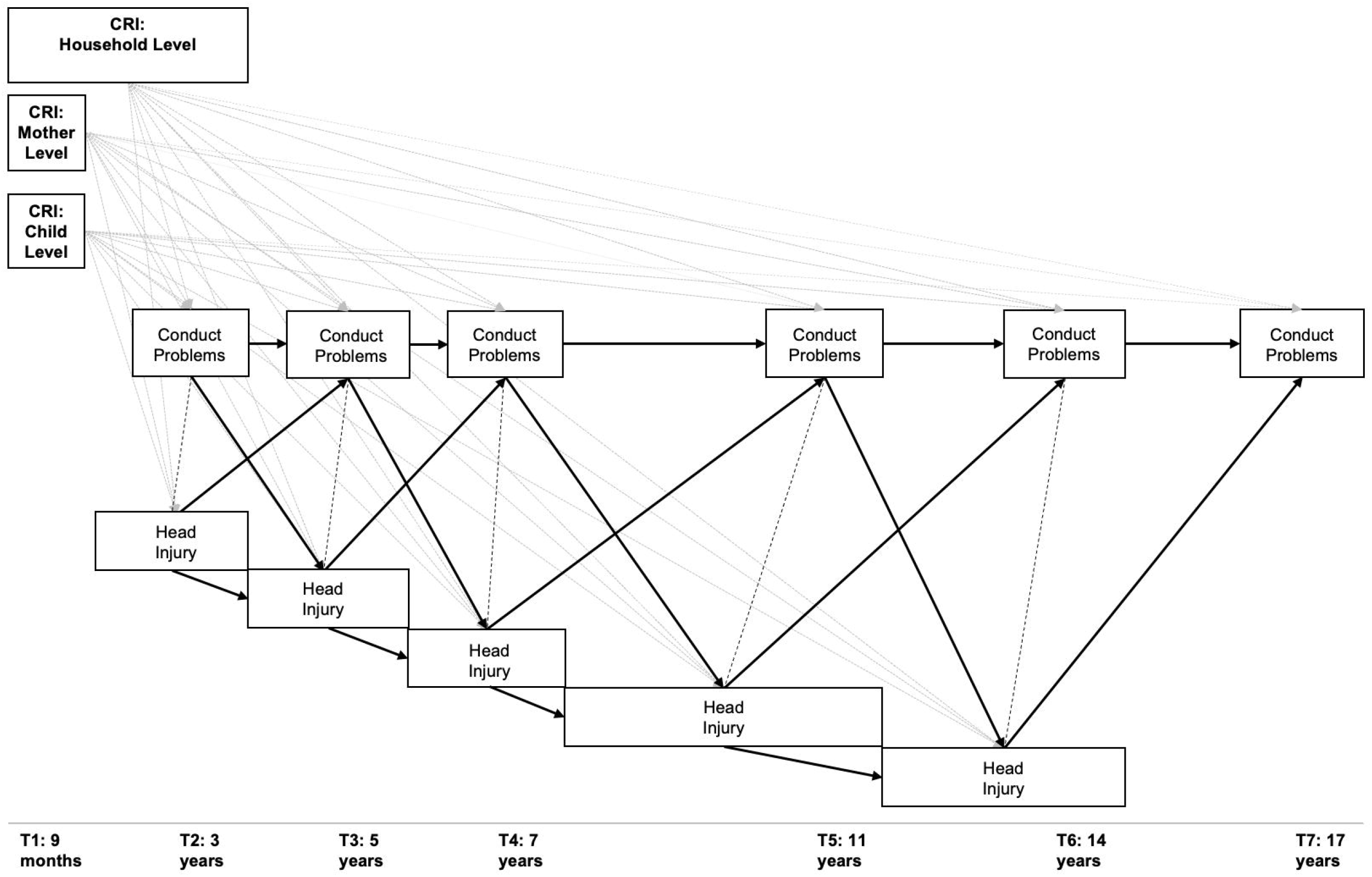
Stylized Illustration of the Structural Equation Model Implemented in This Study. This figure shows the cross-lagged path model conducted on conduct problem variables from age 3 (T2) to 17 (T7) and head injury variables from age 3 (T2) to 14 (T6). These are connected by contemporaneous correlations as well as lagged paths to T+1 within and across variables. The three cumulative risk indices (CRI) at the child, mother, and household levels are connected to each head injury and conduct problem variable. Dotted lines represent pathways from CRIs to head injury and conduct problems. Solid lines represent pathways between conduct problem and head injury variables. Dashed lines represent correlations within timepoints. *Note*. At each timepoint head injuries were recorded since the last wave, hence the omission of a break between the head injury variables.

Contemporaneous correlations were added to account for the relationship within-timepoints as is typical of longitudinal modelling [34]. As the correlations were between a binary and continuous variable, Mplus calculated point-biserial (*r*_*pbis*_) correlations.

Satisfactory model fit was evaluated based on Hu and Bentler’s criteria: Tucker-Lewis Index (TLI; satisfactory values ≥ 0.96), the Comparative Fit Index (CFI; satisfactory values ≥ 0.95), and Root Mean Square Error of Approximation (RMSEA; satisfactory values ≤ 0.06) [35]. Where conduct problems (continuous) were the dependent variable, standardised beta values (*ß*) with significance levels were reported. Where head injury (binary) was the dependent variable, the standardised *Z*-value (index of probit regression) was reported. Results were considered significant with α = .05.

#### Data availability

The MCS dataset used in this study is available via the UK Data Service. The Mplus output for the direct and indirect effects as well as the code needed to create the CRI variables can be accessed via Pure.

## Results

### Participants and Demographics

Table 1 provides a summary and comparison of sample characteristics between the excluded and analytical samples. The analytical sample differed significantly from the excluded sample on all variables, though these effects were small for the head injury variables (Cramer’s *V*=.03 to .08) and medium for child sex (*d* =.50). A further breakdown of the head injury variable can be seen in Supplementary Table 2.

**Table 1.**
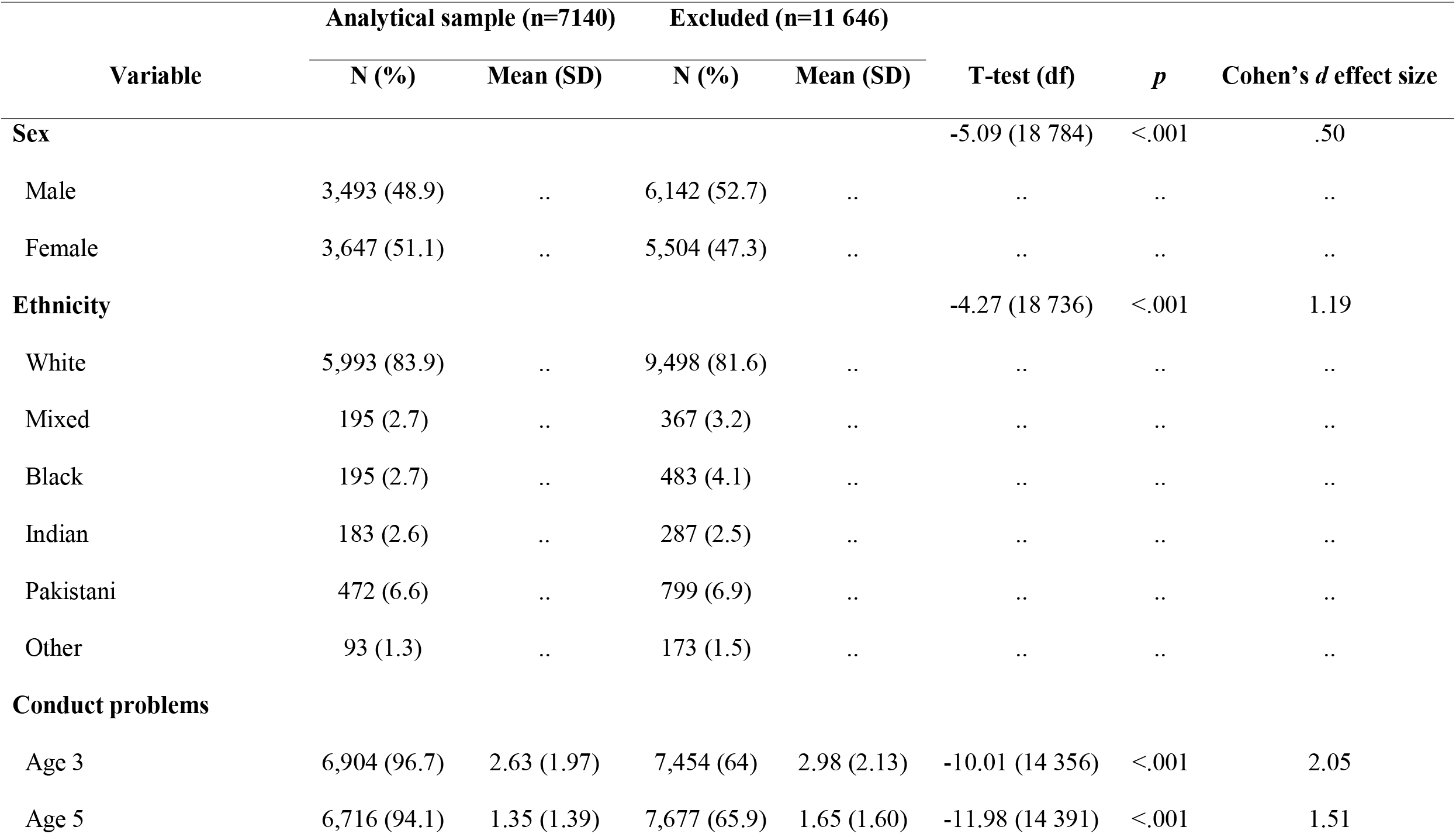
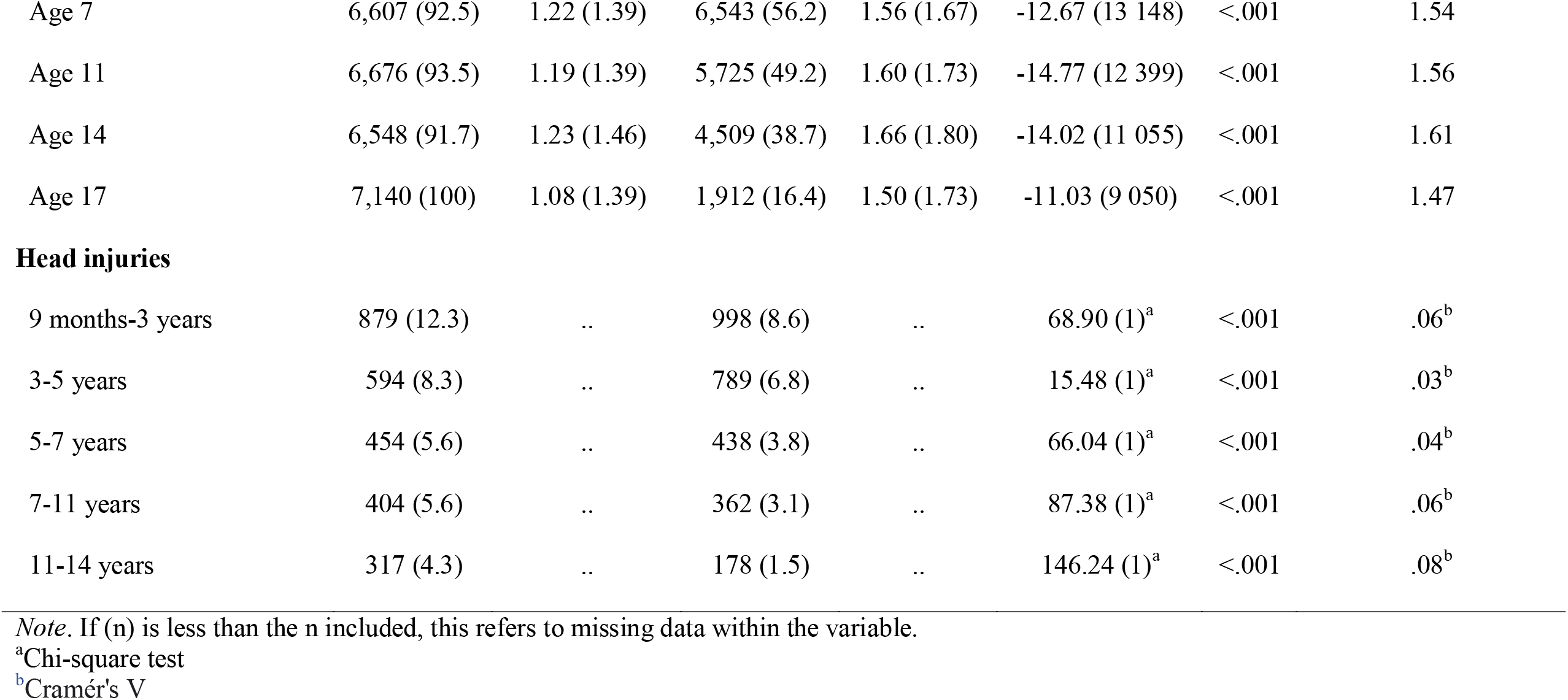
Characteristics of and Differences Between the Analytical Sample (n=7140) and Excluded Sample (n=11646)

| Variable | Analytical sample (n=7140) |  | Excluded (n=11 646) |  | T-test (df) | *p* | Cohen's *d* effect size |
| --- | --- | --- | --- | --- | --- | --- | --- |
|  | N (%) | Mean (SD) | N (%) | Mean (SD) |  |  |  |
| **Sex** |  |  |  |  | -5.09 (18 784) | <.001 | .50 |
| Male | 3,493 (48.9) | .. | 6,142 (52.7) | .. | .. | .. | .. |
| Female | 3,647 (51.1) | .. | 5,504 (47.3) | .. | .. | .. | .. |
| **Ethnicity** |  |  |  |  | -4.27 (18 736) | <.001 | 1.19 |
| White | 5,993 (83.9) | .. | 9,498 (81.6) | .. | .. | .. | .. |
| Mixed | 195 (2.7) | .. | 367 (3.2) | .. | .. | .. | .. |
| Black | 195 (2.7) | .. | 483 (4.1) | .. | .. | .. | .. |
| Indian | 183 (2.6) | .. | 287 (2.5) | .. | .. | .. | .. |
| Pakistani | 472 (6.6) | .. | 799 (6.9) | .. | .. | .. | .. |
| Other | 93 (1.3) | .. | 173 (1.5) | .. | .. | .. | .. |
| **Conduct problems** |  |  |  |  |  |  |  |
| Age 3 | 6,904 (96.7) | 2.63 (1.97) | 7,454 (64) | 2.98 (2.13) | -10.01 (14 356) | <.001 | 2.05 |
| Age 5 | 6,716 (94.1) | 1.35 (1.39) | 7,677 (65.9) | 1.65 (1.60) | -11.98 (14 391) | <.001 | 1.51 |
| Age 7 | 6,607 (92.5) | 1.22 (1.39) | 6,543 (56.2) | 1.56 (1.67) | -12.67 (13 148) | <.001 | 1.54 |
| Age 11 | 6,676 (93.5) | 1.19 (1.39) | 5,725 (49.2) | 1.60 (1.73) | -14.77 (12 399) | <.001 | 1.56 |
| Age 14 | 6,548 (91.7) | 1.23 (1.46) | 4,509 (38.7) | 1.66 (1.80) | -14.02 (11 055) | <.001 | 1.61 |
| Age 17 | 7,140 (100) | 1.08 (1.39) | 1,912 (16.4) | 1.50 (1.73) | -11.03 (9 050) | <.001 | 1.47 |
| Head injuries |  |  |  |  |  |  |  |
| 9 months-3 years | 879 (12.3) | .. | 998 (8.6) | .. | 68.90 (1)<sup>a</sup> | <.001 | .06<sup>b</sup> |
| 3-5 years | 594 (8.3) | .. | 789 (6.8) | .. | 15.48 (1)<sup>a</sup> | <.001 | .03<sup>b</sup> |
| 5-7 years | 454 (5.6) | .. | 438 (3.8) | .. | 66.04 (1)<sup>a</sup> | <.001 | .04<sup>b</sup> |
| 7-11 years | 404 (5.6) | .. | 362 (3.1) | .. | 87.38 (1)<sup>a</sup> | <.001 | .06<sup>b</sup> |
| 11-14 years | 317 (4.3) | .. | 178 (1.5) | .. | 146.24 (1)<sup>a</sup> | <.001 | .08<sup>b</sup> |

*Note.* If (n) is less than the n included, this refers to missing data within the variable.
<sup>a</sup>Chi-square test
<sup>b</sup>Cramér's V

### Association Between Head Injury and Conduct Problems Across Development

The cross-lagged path model estimated to answer the research questions showed good fit with the data (χ^2^(32) = 533.01; *p* < .001; RMSEA = .05 [.04, .05]; CFI = .96; TLI = .88).

The contemporaneous correlations between head injury and conduct problems were small, positive, (*r*_*pbis*_ < .14) and significant (*p* < .05) for all ages except ages 5 (*r*_*pbis*_ = -.01; *SE* = .03; *p* = .753) and 7 (*r*_*pbis*_ = .03; *SE* = .03; *p* = .471).

Head injury at each time point had significant direct (Figure 2, Supplementary Table 3) and indirect effects (Figure 2, Supplementary Table 4) for an increased likelihood of subsequent head injury, as did conduct problems for increased subsequent conduct problems (Figure 2, Supplementary Tables 3 and 4).

**Figure 2.**
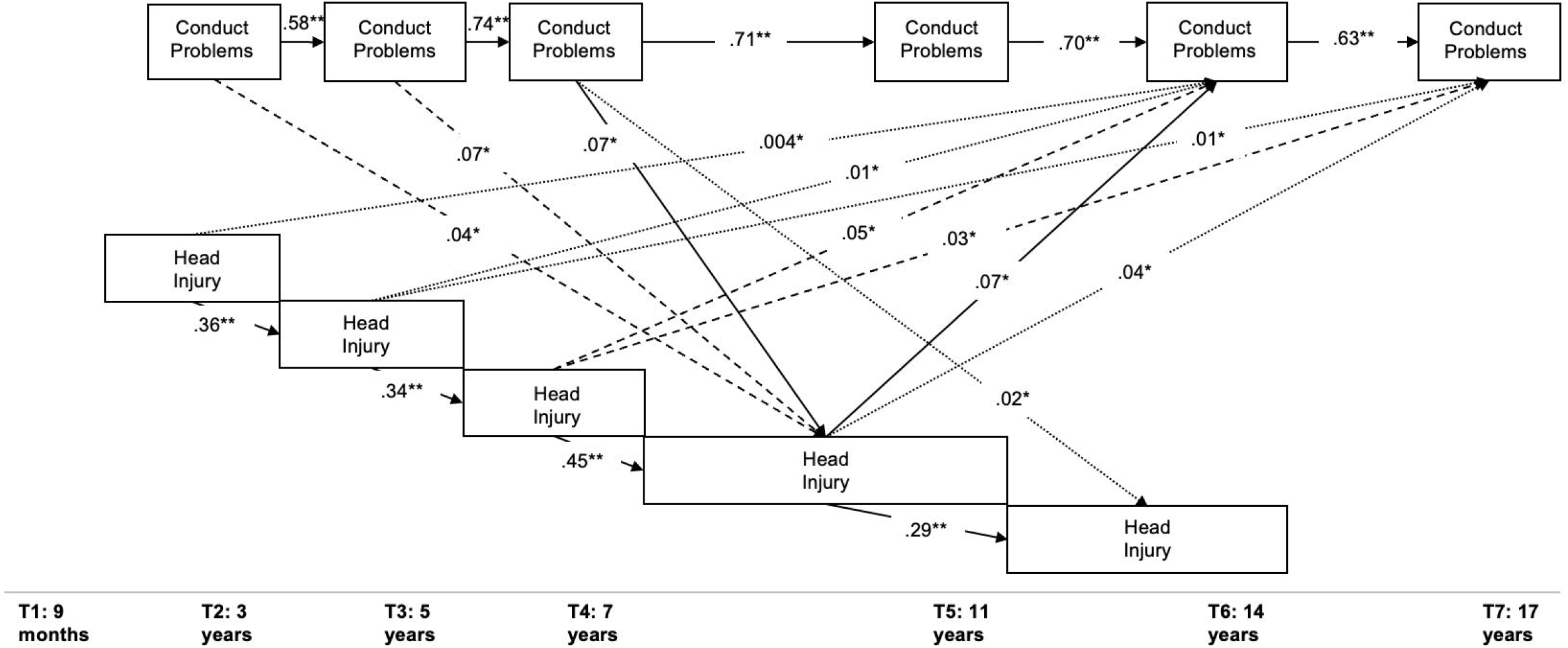
The Direct and Indirect Effects Within and Between Conduct Problems and Head Injury From Ages 3 to 17. This figure shows the significant direct effects (solid lines) within the head injury and conduct problem variables and between them as well as the significant total indirect (dashed lines) and specific indirect (dotted lines) effects. All indirect effects from head injury to later head injury variables (T+1 onwards) and from conduct problems to later conduct problem variables were significant but omitted for clarity. Only significant pathways are shown to *p*<.05 (*) and *p*<.001 (**).

Head injury at ages 7 to 11 had a direct effect for increased conduct problems at age 14 (*ß* = .07; *SE* = .03; 95% CI, .01-.12). Head injuries at ages 5 to 7 had significant total indirect effects on subsequent conduct problems at ages 14 and 17 14 (*ß* = .05; *SE* = .02; 95% CI, .01-.09; *ß* = .03; *SE* = .01; 95% CI, .004-.05), respectively). Further, head injuries at ages 9 months to 3 and 3 to 5 had significant individual indirect effects linked to greater conduct problems at age 14 (*ß* = .004; *SE* = .002; 95% CI, .001-.01; *ß* = .01; *SE* = .004; 95% CI, .002-.02 respectively). Head injuries at ages 3 to 5 and 7 to 11 had significant individual indirect effects linked to greater conduct problems at age 17 (*ß* = .01; *SE* = .003; 95% CI, .001-.01; *ß* = .04; *SE* = .02; 95% CI, .01-.07, respectively). See Figure 2 for visualisation.

Conduct problems at age 7 had a direct effect for an increased likelihood of head injury at age 11 (*Z* = .07; *SE* = .03; 95% CI, .01-.12). There were significant total indirect effects from conduct problems at ages 3 and 5 for an increased likelihood of head injuries at ages 7 to 11 (*Z* = .04; *SE* = .01; 95% CI, .01-.07; *Z* = .07; *SE* = .02; 95% CI, .02-.11, respectively). Significant individual indirect effects were identified from conduct problems at age 7 for an increased likelihood of head injuries at ages 11 to 14 (*Z* = .02; *SE* = .01; 95% CI, .001-.04). See Figure 2 for visualisation.

### The Influence of Child, Mother and Household-level Demographic Risk Factors

Child-level cumulative risk had a significant direct effect for increased conduct problems at ages 3, 5, and 7 (Table 2, Supplementary Figure 2). Mother-level cumulative risk had a significant direct effect for increased conduct problems at ages 3, 5, 7, 14, and 17 (Table 2, Supplementary Figure 3). Household-level cumulative risk had a significant direct effect for increased conduct problems at ages 3, 11, and 17 (Table 2, Supplementary Figure 4). All three CRIs had significant total indirect effects for increased conduct problems from ages 5 to 17 (Table 2, Supplementary Figures 2, 3, and 4).

**Table 2.**
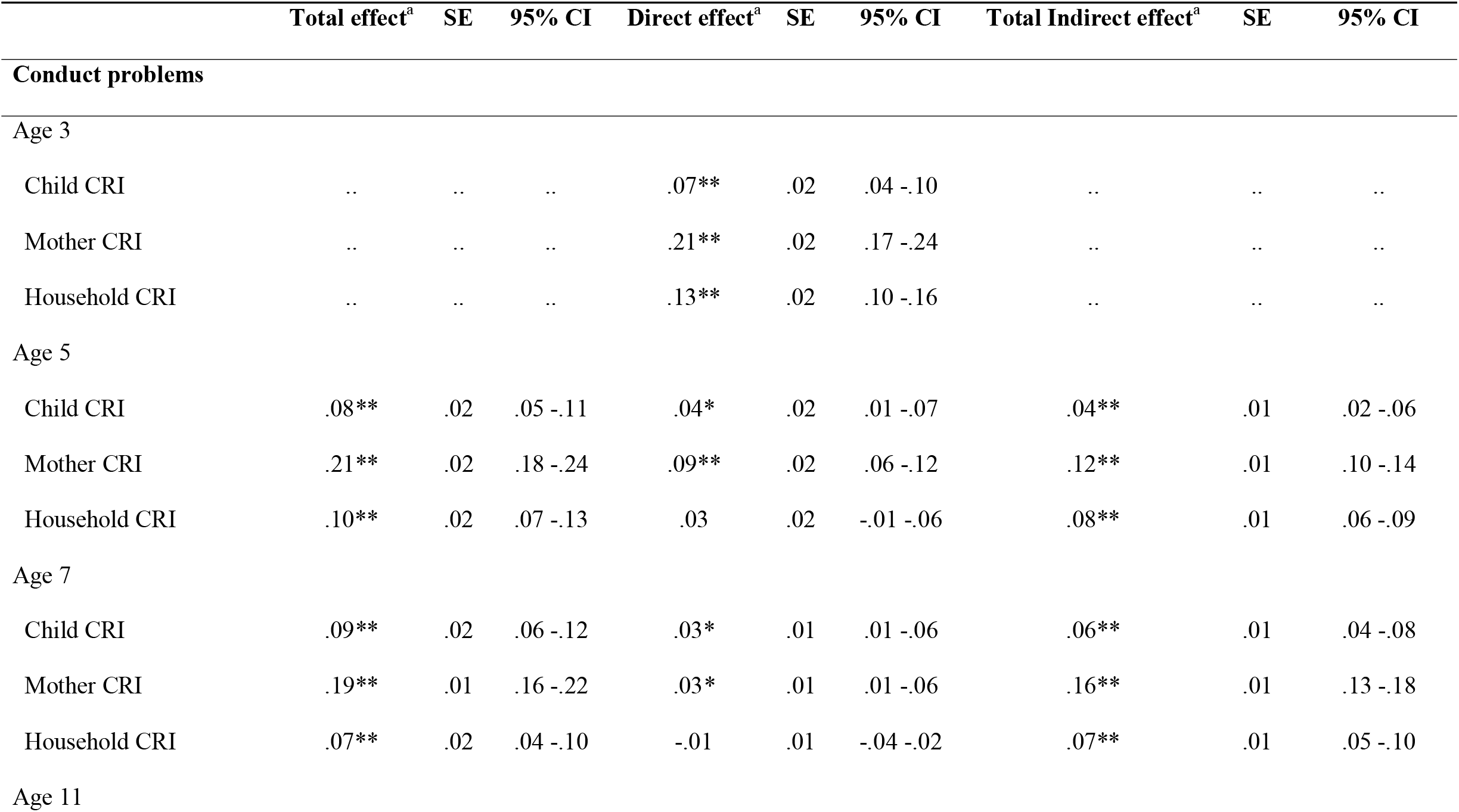
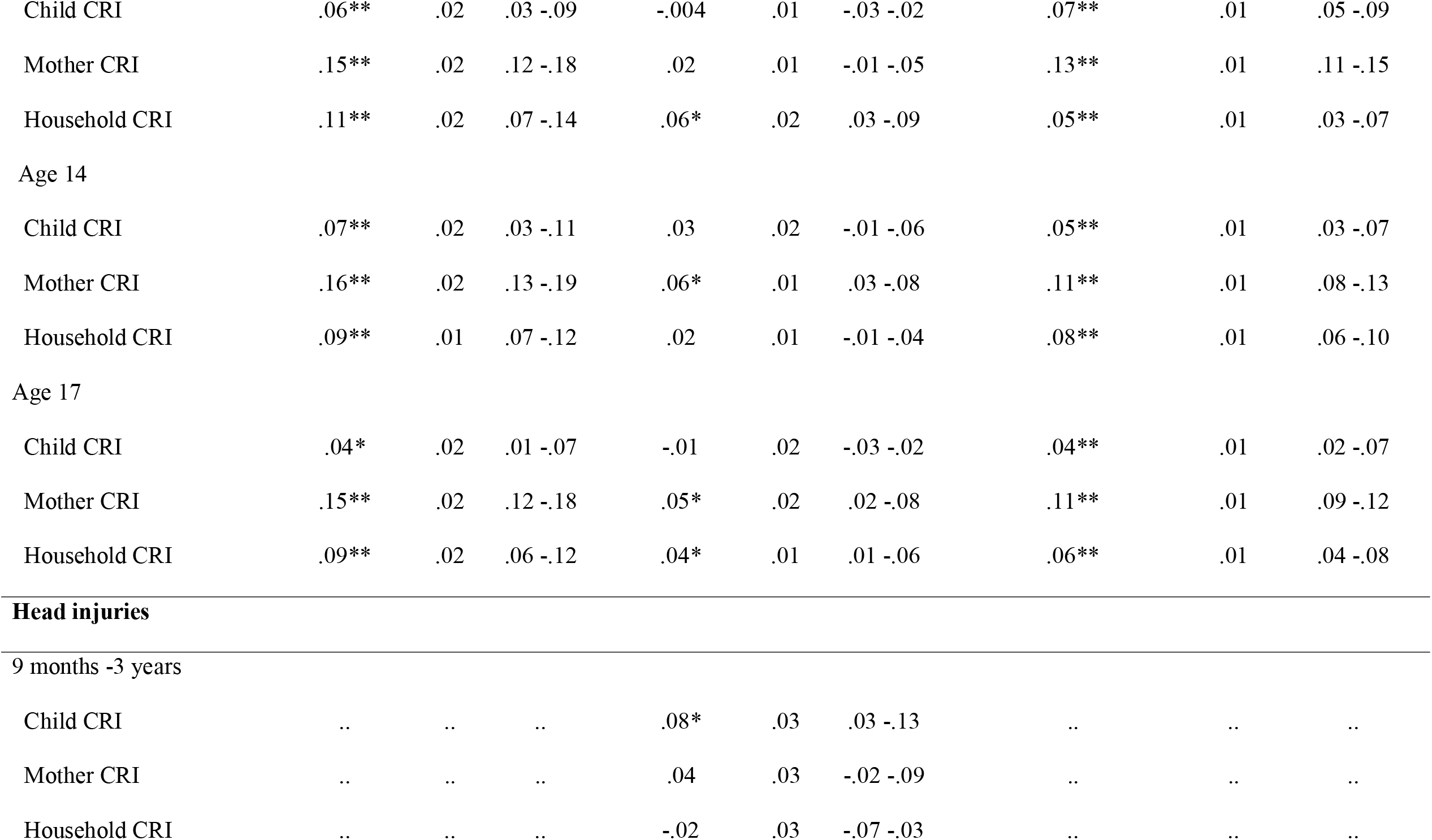
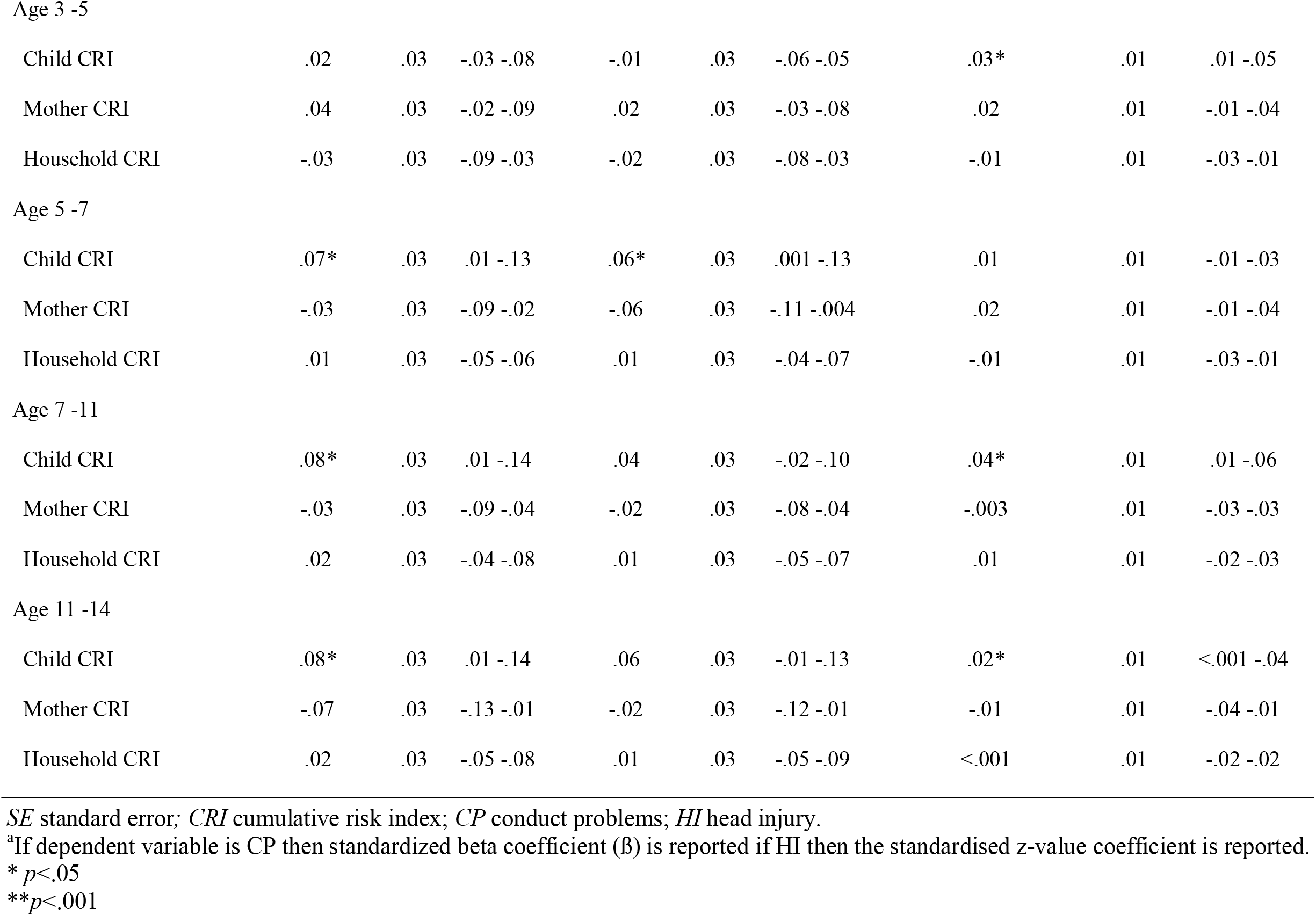
The Total, Direct, and Total Indirect Effects of the Child, Mother and Household CRIs on Conduct Problems and Head Injury.

Only the child-level cumulative risk had a significant direct effect for an increased likelihood of head injuries from 9 months to 3 years, and from 5 to 7 years (see Table 2, Supplementary Figure 2). Total indirect effects were significant only at the child-level for head injuries sustained at ages 3 to 5, 7 to 11, and 11 to 14 (Table 2, Supplementary Figure 2).

Significant individual indirect effects were also present from the child-level to head injuries sustained at ages 5 to 7 (*Z* = .01; *SE*=.004; 95% CI, .003-.02), and the mother level for ages 7 to 11 (*Z* = .01; *SE*=.003; 95% CI, .001-.01).

## Discussion

The aim of this study was to identify if there were bidirectional associations between conduct problems and head injuries in a UK population between the ages of 3 and 17 years, while controlling for salient demographic risk factors. The results showed that higher levels of conduct problems at age 7 promoted an increased likelihood of head injury between the ages of 7 to 11 whilst a head injury sustained between the ages of 7 and 11 promoted increased conduct problems at age 14. Thus, this study shows a longitudinal, bidirectional relationship between head injuries and conduct problems during a sensitive period between the ages of 7 and 11 years. Further, the bidirectional relationship between head injury and conduct problems exists over and above the effects of salient child, mother, and household demographic risk factors.

These results provide further evidence that childhood head injuries are associated with increased levels of conduct problems [2]. However, it elaborates on the previous literature by suggesting that this relationship is bidirectional and that conduct problems also promote head injuries during the sensitive period of 7 to 11 years. This was only previously identified when there was a co-morbid diagnosis of ADHD [11] or in a young adult population [12]. This clarifies results shown by Brandt and colleagues [3] whilst controlling for salient demographic risk. Thus, the current study provides novel insight into a potential bidirectional association between head injury and conduct problems that warrants further investigation.

In line with existing literature, child, mother, and household demographic risks all had direct and indirect effects for increased conduct problems over the course of development (from 3 to 17 years) [22,25-27]. However and surprisingly, the mother and household risks were found to play no direct role in promoting head injuries during childhood (from age 9 months). Direct risk instead lied solely at the level of the child with all but one of these risk factors (male sex) being themselves socially stratified.

### Strengths and limitations

The key strength of the current study is its use of a large birth cohort dataset which enabled the statistical unpacking of the complex relationships linking conduct problems to head injuries and vice versa over time. Another strength is the comprehensive inclusion of all head injuries (any that resulted in a parent seeking medical aid), which increases the ecological validity to the findings.

A limitation of this paper is the stringent inclusion criteria for participants, which while reducing bias from confounding variables, limited the generalizability of the findings to the general UK population. The analytical sample differed from the total sample on demographics including ethnicity. Therefore, the results may not reflect the ethnic diversity within the UK population meaning that these results must be read with caution whilst directing future research with non-white children and families in the UK. The analytical sample also had significantly lower mean conduct problems than the total sample, suggesting that the sample may not be representative of conduct problems presented in the general UK population. This difference may be due to the exclusion of those with a diagnosis of ADHD or epilepsy because conduct problems are highly comorbid with both [36] (thus excluding the latter two risks increasing the former). Hence, the current findings do not generalise to children with a diagnosis of epilepsy or ADHD.

Parent-report for both head injuries and conduct problems might be considered a limitation. Though this addresses the limitations of previous head injury research whereby self-report is likely to inhibit accuracy (i.e. due to infantile amnesia) [37], it could introduce a social desirability bias. Therefore, this research (as with all research using parent measures) requires smaller-scale follow-up using more objective measures, such as clinical records.

### Implications

Parents and teachers of children with high levels of conduct problems between 7 and 11 years should be made more aware that there is an increased risk of head injury and should consider additional safety precautions. This is particularly important because the increased risk for head injuries during this period appears to pose a further risk for a subsequent increase in conduct problems all the way through to age 14.

Parents and primary schools may also consider limiting or prohibiting contact sports in children under 11 years old where there is the potential to sustain a head injury (e.g. within certain forms of contact sport) [38]. Helmet usage is also encouraged when riding a bike with interventions encouraged to promote helmet use [39]. Such interventions will lower the risk of sustaining future head injuries and also the risk of developing conduct problems through to age 14.

### Conclusions

The results of this study suggest a sensitive period between the ages of 7 and 11 where conduct problems and head injuries are risk factors for one another with consequences for interventions that run both before and during this period.

## Data Availability

https://beta.ukdataservice.ac.uk/datacatalogue/series/series?id=2000031#!/access-data

## Statements and Declarations

## Acknowledgments

HC was funded by the Economic and Social Research Council South Coast Doctoral Training Partnership (Grant Number ES/P000673/1). We are grateful to the Centre for Longitudinal Studies (CLS), UCL Social Research Institute, for the use of the MCS dataset and to the UK Data Service for making them available. Neither CLS nor the UK Data Service bear any responsibility for the analysis or interpretation of these data.

