## Supplement for "The Bidirectional Relationship Between Head Injuries and Conduct Problems: Longitudinal Modelling of a Population-Based Birth Cohort Study"

**Supplementary material**

### **Supplementary tables**

### **Table 1. Salient Demographic Risk Factors by Ecological Level (Child, Mother, and Household) Measured via Cumulative Risk Indices**

|  | **No. (%) with data^a^** | **Mean (SD)** | **Cut-off for high-risk** | **No. (%) high-risk** |
| --- | --- | --- | --- | --- |
| **Child-level cumulative risk index** |  |  |  |  |
| Sex | 7,140 (100) |  | Male sex | 3,493 (48.9) |
| Male | 3,493 (48.9) |  |  |  |
| Female | 3,647 (51.1) |  |  |  |
| Birth weight | 7,134 (99.9) | 3.79 (0.58) | < 2.5kg | 449 (6.3) |
| Gestation | 7,087 (99.2) | 276.33 (13.48) | <= 252 days gestation | 387 (5.4) |
| Pregnancy smoking status | 7,130 (99.9) |  | >= 1 cigarette smoked | 1,257 (17.6) |
| Smoked | 1,257 (17.6) |  |  |  |
| Not smoked | 5,883 (82.5) |  |  |  |
| Pregnancy alcohol consumption | 7,140 (100) |  | Any alcohol consumption | 2,305 (32.3) |
| Everyday | 28 (0.4) |  |  |  |
| 5-6 times per week | 16 (0.2) |  |  |  |
| 3-4 times per week | 102 (1.4) |  |  |  |
| 1-2 times per week | 545 (7.6) |  |  |  |
| 1-2 times per month | 549 (7.7) |  |  |  |
| Less than once a month | 1,065 (14.9) |  |  |  |
| Never | 4,835 (67.7) |  |  |  |
| Child-level cumulative risk index | 7,087 (99.3) |  | Percentage encountering:  4+ risks  3 risks  2 risks  1 risk  No risk | 85 (1.2)  363 (5.1)  1,667 (23.3)  3,122 (43.7)  1,903 (26.7) |
| **Mother-level cumulative risk index** |  |  |  |  |
| Age at pregnancy | 7,140 (100) | 29.38 (5.67) | <18 years old | 102 (1.4) |
| Highest attained level of education | 7,140 (100) |  | No high-school qualification | 947 (13.3) |
| No education | 947 (13.3) |  |  |  |
| NVQ 1 equivalent | 628 (8.8) |  |  |  |
| NVQ 2 equivalent | 2,325 (32.6) |  |  |  |
| NVQ 3 equivalent | 769 (10.8) |  |  |  |
| NVQ 4 equivalent | 1,967 (27.5) |  |  |  |
| NVQ 5 equivalent | 329 (4.6) |  |  |  |
| Overseas | 175 (2.5) |  |  |  |
| Employment status | 7,140 (100) |  | Unemployed | 3,270 (45.8) |
| Employed | 3,870 (54.2) |  |  |  |
| Unemployed | 3,270 (45.8) |  |  |  |
| Attachment | 6,955 (97.4) | 24.39 (3.30) | =< 22 on Condon Maternal Attachment Scale | 1,762 (24.7) |
| Psychological distress | 7,138 (100) | 1.51 (1.64) | >= 4 on Rutter Malaise Inventory | 833 (11.7) |
| Mother-level cumulative risk index | 6,954 (97.4) |  | Percentage encountering:  4+ risks  3 risks  2 risks  1 risk  No risk | 61 (0.8)  391 (5.6)  1,251 (17.5)  2,739 (38.4)  2,506 (35.1) |
| **Household-level cumulative risk index** |  |  |  |  |
| Parents in household | 7,140 (100) |  | Single parent | 828 (11.6) |
| Single parent | 828 (11.6) |  |  |  |
| Two parents | 6,312 (88.4) |  |  |  |
| Household income | 7,129 (99.8) |  | Below 60% poverty indicator | 1,988 (27.8) |
| Above 60% poverty indicator | 5,141 (72.1) |  |  |  |
| Below 60% poverty indicator | 1,988 (27.9) |  |  |  |
| Household crowding | 7,134 (99.9) |  | Fewer rooms than people^b^ | 690 (9.7) |
| Overcrowded | 690 (9.7) |  |  |  |
| Not overcrowded | 6,444 (90.3) |  |  |  |
| Highest occupational status in household | 7,140 (100) |  | Semi-skilled or lower | 1,595 (22.3) |
| Unemployed | 1,130 (15.8) |  |  |  |
| Semi-routine or less | 465 (6.5) |  |  |  |
| Low supervisor or technical | 372 (5.2) |  |  |  |
| Self-employed | 420 (5.9) |  |  |  |
| Intermediate | 1,029 (14.4) |  |  |  |
| Managerial | 3,724 (52.2) |  |  |  |
| Early Home Learning Environment | 7,140 (100) | 25.19 (7.58) | Bottom quartile | 1,900 (26.6) |
| Household-level cumulative risk index | 7,129 (99.8) |  | Percentage encountering:  4+ risks  3 risks  2 risks  1 risk  No risk | 49 (0.7)  428 (6.0)  1,385 (19.4)  2,749 (38.5)  2,529 (35.4) |

^a^No. less than 7140 indicates missing data in the variable.

^b^Excluding bathrooms and hallways.

### **Table 2. A Breakdown of Head Injury Reporting per Timepoint**

|  | | **Head injury** | | |
| --- | --- | --- | --- | --- |
| **MCS Timepoint** | **Age (years)** | **No. (%) who sustained any head injury** | **No. (%) who sustained a bang on the head** | **No. (%) who sustained a head injury with LOC** |
| 2 | 3 | 879 (12.3) | 835 (11.7) | 44 (0.6) |
| 3 | 5 | 594 (8.3) | 576 (8.1) | 22 (0.3) |
| 4 | 7 | 454 (6.4) | 434 (6.1) | 27 (0.4) |
| 5 | 11 | 404 (5.7) | 356 (5.0) | 55 (0.8) |
| 6 | 14 | 317 (4.4) | 241 (3.4) | 85 (1.2) |
| 7 | 17 | .. | .. | .. |

*MCS* Millennium Cohort Study; *LOC* loss of consciousness

### **Table 3. The Direct Effects of Conduct Problems and Head Injury Over Time**

| **Timepoints** | **Conduct problems -> conduct problems** | | |  | **Head injury -> conduct problems** | | |  | **Head injury**  **-> head injury** | | | |  | **Conduct problems –> head injury** | | |
| --- | --- | --- | --- | --- | --- | --- | --- | --- | --- | --- | --- | --- | --- | --- | --- | --- |
|  | ***ß*** | **SE** | **95% CI** |  | ***ß*** | **SE** | **95% CI** |  | | ***Z*** | **SE** | **95% CI** |  | ***Z*** | **SE** | **95% CI** |
| 2 –> 3 | .58 | .01** | .56 to .60 |  | .03 | .03 | -.02 to .08 |  | | .36** | .04 | .29 to .43 |  | .01 | .03 | -.04 to .06 |
| 3 –> 4 | .74 | .01** | .72 to .76 |  | .001 | .02 | -.05 to .05 |  | | .34** | .04 | .27 to .42 |  | .04 | .03 | -.02 to .09 |
| 4 –> 5 | .71 | .01** | .69 to .72 |  | .03 | .02 | -.02 to .08 |  | | .45** | .04 | .37 to .53 |  | .07* | .03 | .01 to .12 |
| 5 –> 6 | .70 | .01** | .68 to .72 |  | .07* | .03 | .01 to .12 |  | | .29** | .05 | .19 to .38 |  | -.04 | .04 | -.12 to .03 |
| 6 –> 7 | .63 | .01** | .62 to .65 |  | -.03 | .02 | -.08 to .02 |  | | .. | .. | .. |  | .. | .. | .. |

*SE* standard error

* *p*<.05

***p*<.001

### **Table 4. Total Indirect Effects of Conduct Problems and Head Injury on One Another Over Time**

| **Total indirect effect**^a^ | ***ß* or *Z****^b^* | **SE** | **95% CI** |
| --- | --- | --- | --- |
| CP2->CP4 | .43** | .01 | .41 to .45 |
| CP2->CP5 | .30** | .01 | .29 to .32 |
| CP2->CP6 | .21** | .01 | .20 to .23 |
| CP2->CP7 | .14** | .01 | .13 to .15 |
| CP3->CP5 | .52** | .01 | .51 to .54 |
| CP3->CP6 | .37** | .01 | .35 to .39 |
| CP3->CP7 | .24** | .01 | .22 to .25 |
| CP4->CP6 | .50** | .01 | .48 to .52 |
| CP4->CP7 | .32** | .01 | .30 to .33 |
| CP5->CP7 | .44** | .01 | .43 to .46 |
| HI2–>CP4 | .02 | .02 | -.02 to .06 |
| HI2–>CP5 | .02 | .01 | -.01 to .05 |
| HI2–>CP6 | .02 | .01 | -.003 to .04 |
| HI2–>CP7 | .01 | .01 | -.002 to .02 |
| HI3–>CP5 | .01 | .02 | -.02 to .05 |
| HI3–>CP6 | .02 | .01 | -.01 to .04 |
| HI4–>CP6 | .05* | .02 | .01 to .09 |
| HI3–>CP7 | .01 | .01 | -.01 to .03 |
| HI4–>CP7 | .03* | .01 | .004 to .05 |
| HI5–>CP7 | .03 | .02 | -.001 to .07 |
| HI2->HI4 | .13** | .02 | .09 to .16 |
| HI2->HI5 | .06** | .01 | .04 to .08 |
| HI2->HI6 | .02** | .004 | .01 to .02 |
| HI3->HI5 | .15** | .03 | .11 to .20 |
| HI3->HI6 | .04** | .01 | .02 to .06 |
| HI4->HI6 | .13** | .03 | .08 to .18 |
| CP2–>HI4 | .03 | .02 | -.01 to .06 |
| CP2–>HI5 | .04* | .01 | .01 to .07 |
| CP2–>HI6 | -.002 | .01 | -.02 to .02 |
| CP3–>HI5 | .07* | .02 | .02 to .11 |
| CP3–>HI6 | -.004 | .02 | -.04 to .04 |
| CP4->HI6 | -.01 | .03 | -.06 to .04 |
| HI2->HI3->HI4->HI5->CP6 ^c^ | .004* | .002 | .001 to .01 |
| HI3->HI4->HI5->CP6 ^c^ | .01* | .004 | .002 to .02 |
| HI3->HI4->HI5->CP6->CP7 ^c^ | .01* | .003 | .001 to .01 |
| HI5->CP6->CP7 | .04* | .02 | .01 to .07 |
| CP4->HI5->HI6 ^c^ | .02* | .01 | .001 to .04 |

*SE* standard error*; CP* conduct problems; *HI* head injury.

^a^CPX = conduct problems where X represents the timepoint of measurement (e.g., CP2 = conduct problems at timepoint 2), HIX = head injuries where X represents the timepoint of measurement (e.g., HI2 = head injuries measured at timepoint 2).

^b^If dependent variable is CP then standardized beta coefficient (ß) is reported if HI then the standardised z-value coefficient is reported.

^c^Individual indirect effects.

**p*<.05

***p*<.001

### **Supplementary Figures**

### **Figure 1. A flow chart of the excluded and total analytical sample.**

Enrolled in study at T1 (N=18,786)

Completed T7

(N=10,345)

First-born (N=10,238)

Main respondent biological mother

(N=8,616)

Not first-born child (N=107)

Lost to follow-up

(N=8,441)

Missing SDQ information (N=945)

Completed T7 SDQ

(N=9,293)

Main respondent not the biological mother (N=677)

Diagnosis of ADHD or epilepsy from T1 to T7 (N=1,476)

No history of ADHD or epilepsy (N=7,140)

Final Analytical Sample (N=7,140)

This figure shows the five exclusions made for the current study. It shows the number of participants excluded from the original total sample of N = 18,786 at timepoint 1 (T1) resulting in the final analytical sample of N= 7,140.

### **Figure 2. The Significant Direct and Indirect Effects of Child Level Risk on Conduct Problems and Head Injury.**

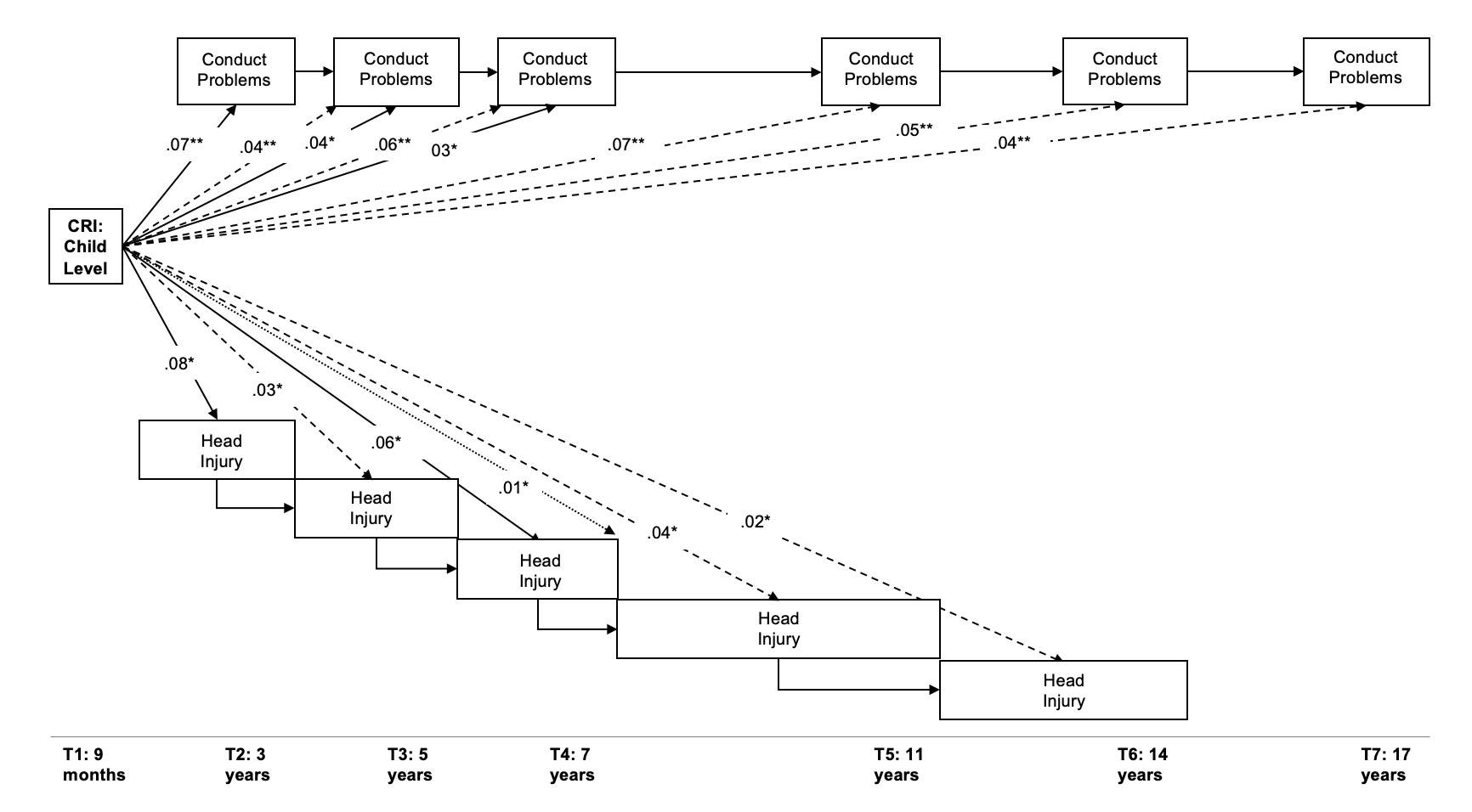

*CRI* cumulative risk index; *T1* timepoint one (same pattern for subsequent timepoints).

This figure shows the significant direct effects from head injury variables to the next wave of head injuries (T+1) and from conduct problems to the next wave of conduct problems. Significant direct effects (solid lines) from the child-level CRI onto conduct problems and head injuries across waves are included. It also shows the total indirect effects (dashed lines) to head injury to conduct problem variables from T3 onwards. One significant specific indirect effect is also shown (dotted line) from the child-level CRI to head injury at T4.

### **Figure 3. The Significant Direct and Indirect Effects of Mother Level Risk on Conduct Problems and Head Injury.**

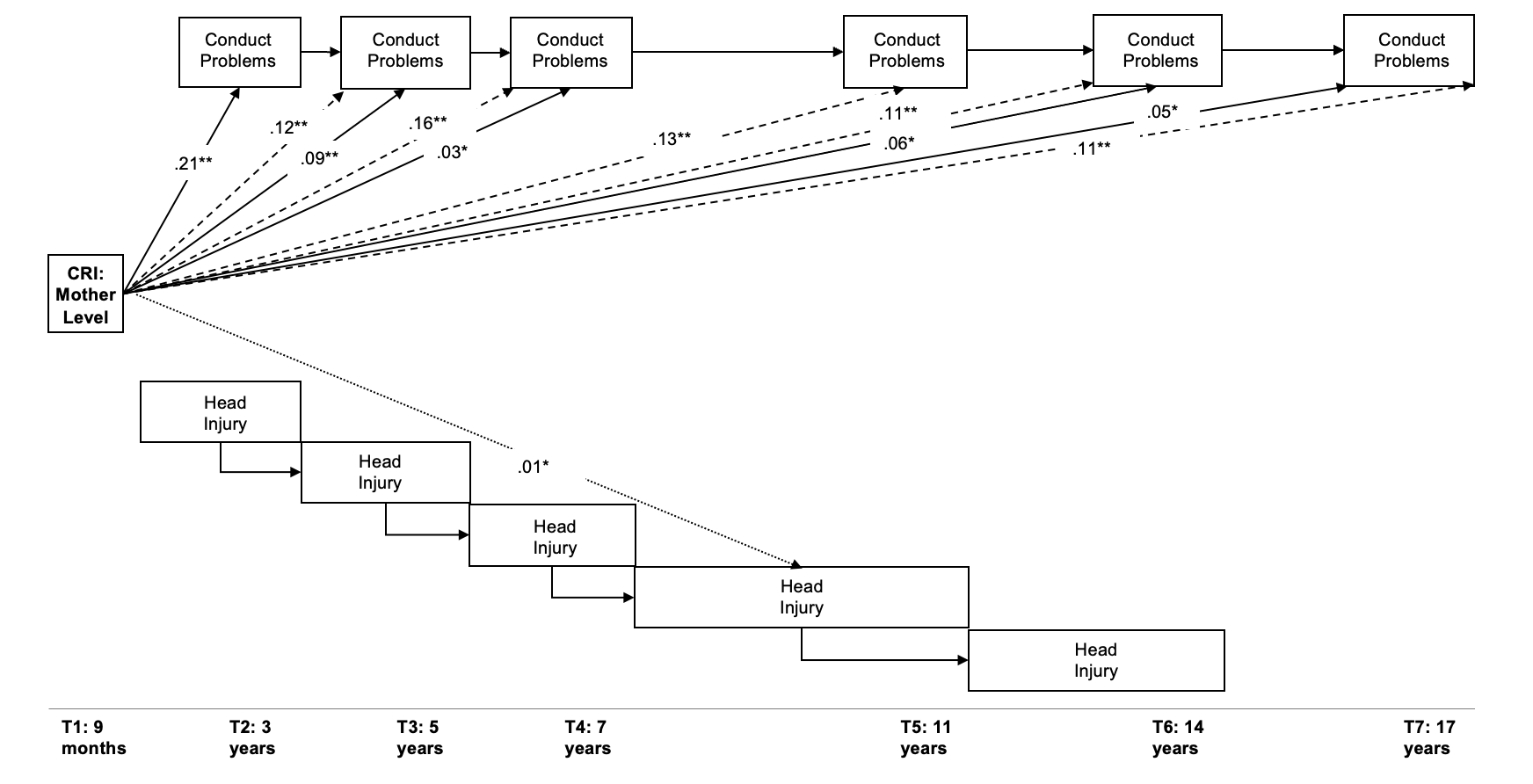

*CRI* cumulative risk index; *T1* timepoint one (same pattern for subsequent timepoints).

This figure shows the significant direct effects from head injury variables to the next wave of head injuries (T+1) and from conduct problems to the next wave of conduct problems. Significant direct effects (solid lines) from the mother-level CRI onto conduct problem variables are included. It also shows the total indirect effects (dashed lines) on conduct problem variables from T3 onwards. Significant specific indirect effects are shown (dotted line) from the mother-level CRI to head injury at T5 and T6.

### **Figure 4. The Significant Direct and Indirect Effects of Household Level Risk on Conduct Problems and Head Injury.**

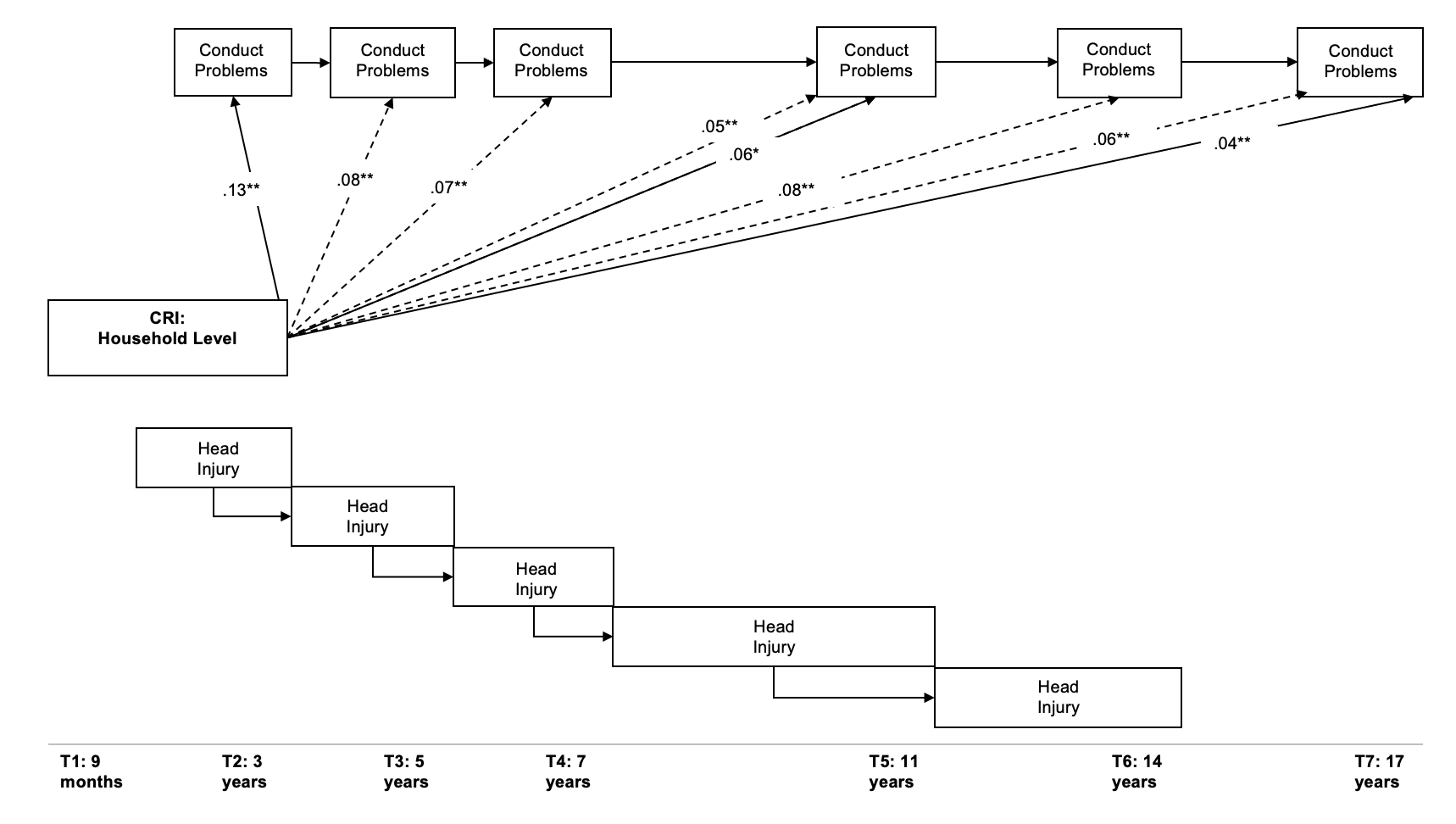

*CRI* cumulative risk index; *T1* timepoint one (same pattern for subsequent timepoints).

This figure shows the significant direct effects from head injury variables to the next wave of head injuries (T+1) and from conduct problems to the next wave of conduct problems. Significant direct effects (solid lines) from the household-level CRI onto conduct problem variables are included. It also shows the total indirect effects (dashed lines) on conduct problem variables from T3 onwards. Significant specific indirect effects are shown (dotted line) from the household-level CRI to head injury at T5 and T6.
